# COVID-19 Infection risk posed by drinking alcohol at restaurants or bars in Japan

**DOI:** 10.1101/2021.07.29.21261358

**Authors:** Junko Kurita, Tamie Sugawara, Yasushi Ohkusa

## Abstract

**Background:** On April 25, 2021, the third state of emergency was declared in Japan. Drinking alcohol at restaurants and bars was banned.

**Object:** We used published data to evaluate drinking ban effects.

**Method:** We bootstrapped the data and evaluated the risk ratio for drinking compared with non-drinking during group dining with similar frequency and numbers of participants.

**Results:** The 99% lower bound of the bootstrapped distribution of the risk ratio was less than one. Therefore the null hypothesis, that incidence in the two styles of group dining were the same, cannot be rejected.

**Discussion and Conclusion:** Results constitute no clear evidence indicating a risk of drinking alcohol in groups. However, further analyses using collected data are necessary. Moreover, risks posed by the number of group diners or their dining frequency must be evaluated.

## Introduction

Under the third declared state of emergency in Japan, restaurants and bars in Tokyo were prohibited from serving alcohol. Although indirect evidence indicates that infections at bars and restaurants might be more common [1], direct evidence, especially for Japan, remains poor. Therefore, alcohol ban policies require stronger evidence to demonstrate their effectiveness.

Under these circumstances, some researchers reported that infection incidence among persons who met together for dining and drinking alcohol with more than two participants had more than twice the infection incidence of persons who had not done so with more than two participants or even once during the prior two weeks [2]. However, because those findings included no separate effects of alcohol consumption, the frequency and number of dining group participants did not implicate alcohol drinking as presenting high risk.

Therefore, they did not analyze group dining and drinking alcohol because of zero incidence. However, given the same scale and frequency of dining, they found the infection incidence among persons who had dined together with more than two participants while drinking alcohol as more than twice that of people dining without alcohol. For the comparison in the present, the numbers of participants and frequency were the same, but some difference might be attributed to alcohol drinking: 7 persons were found to be COVID-19 positive with drinking alcohol, 12 persons were negative with drinking, and 8 persons were negative without drinking. No person was found to be COVID-19 positive without drinking.

Because the incidence among persons without drinking was zero, no case-control study was done. Nevertheless, this finding constitutes information about drinking risks. Therefore, we applied another approach to evaluate this information.

## Methods

For the earlier study [2], a survey was administered during March 30 through June 8 to patients with fever, who had been PCR-tested for COVID-19 infection at two hospitals in Tokyo. Of the 407 patients recruited, only 27 persons had dined together with more than two people more than twice during the prior two weeks.

We examined a non-parametric fully replicated bootstrapping method based on an empirical distribution [3,4,5] for the distribution of {*x*_*i*_ (*i*=1, 2, 3, 4)} representing the number of persons who had tested positive or negative for infection and with or without drinking of alcohol. In addition to the usual bootstrapping, we used a method with special consideration for the case of *x*_*i*_*=*0. These cases were ignored in estimation despite their inclusion of much information. We bootstrapped for the distribution of {*x*_*i*_*+*1(*i*=1,2,3,4)} and obtained {*x*_*i*_*+*1}^*b*^-1(*i*=1, 2, 3, 4), where superscript *b* represents a bootstrapped series.

Based on the *j*-th bootstrapped distribution {*x*_*i*_^b^(*i*=1, 2, 3, 4)}^*j*^, one can obtain risk ratio *R*_,*j*_*, which represents the relative risk posed by drinking. We repeated this procedure one million times, thereby obtaining one million bootstrapped *R*_,*j*_*. We sorted these variables. The duration from *R*_*i*,500_* to *R*_*i*,99500_* is expected to be 99% CI of *R*\*.

In addition to these analyses, we also performed control of the background outbreak situation. If one group was limited to the high epidemic period, then risk of infection was probably higher than in another group, irrespective of drinking. Therefore, the background situation must be adjusted if the period when an observed group was not balanced.

For this study period, the third state of emergency was declared from April 25 through June 20, 2021. For this countermeasure, drinking alcohol at restaurants and bars was banned. From June 21, drinking alcohol at restaurants or bars alone or with one other person was allowed, but meetings with more than two participants continued to be banned. Therefore, a dining or drinking group including more than two participants meeting more than twice during the prior two weeks is assumed to have met before the third state of emergency. Conversely, the non-drinking group can have met at any time during period. Therefore, the background situations of the outbreak might be different. We adjusted the results such that the infection incidence of the drinking group was divided by the difference in prevalence between that before the state of emergency and the entire study period. We adopted 1% as the significance level.

## Results

The sorted *R*_,*j*_*is shown in Figure 1. Of course, about 75% of bootstrapped incidence of drinking was zero. the risk ratio was infinity in this case. However, *R* _,*1*_* was clearly less than one, indicating some probability that group dining with drinking is less risky than group dining without drinking. Actually, its probability was estimated as 2.25%. Because this probability is higher than half of the significance level of 0.5%, the null hypothesis, that the infection incidences of the two types of meeting were not different, cannot be rejected.

**Figure 1:**
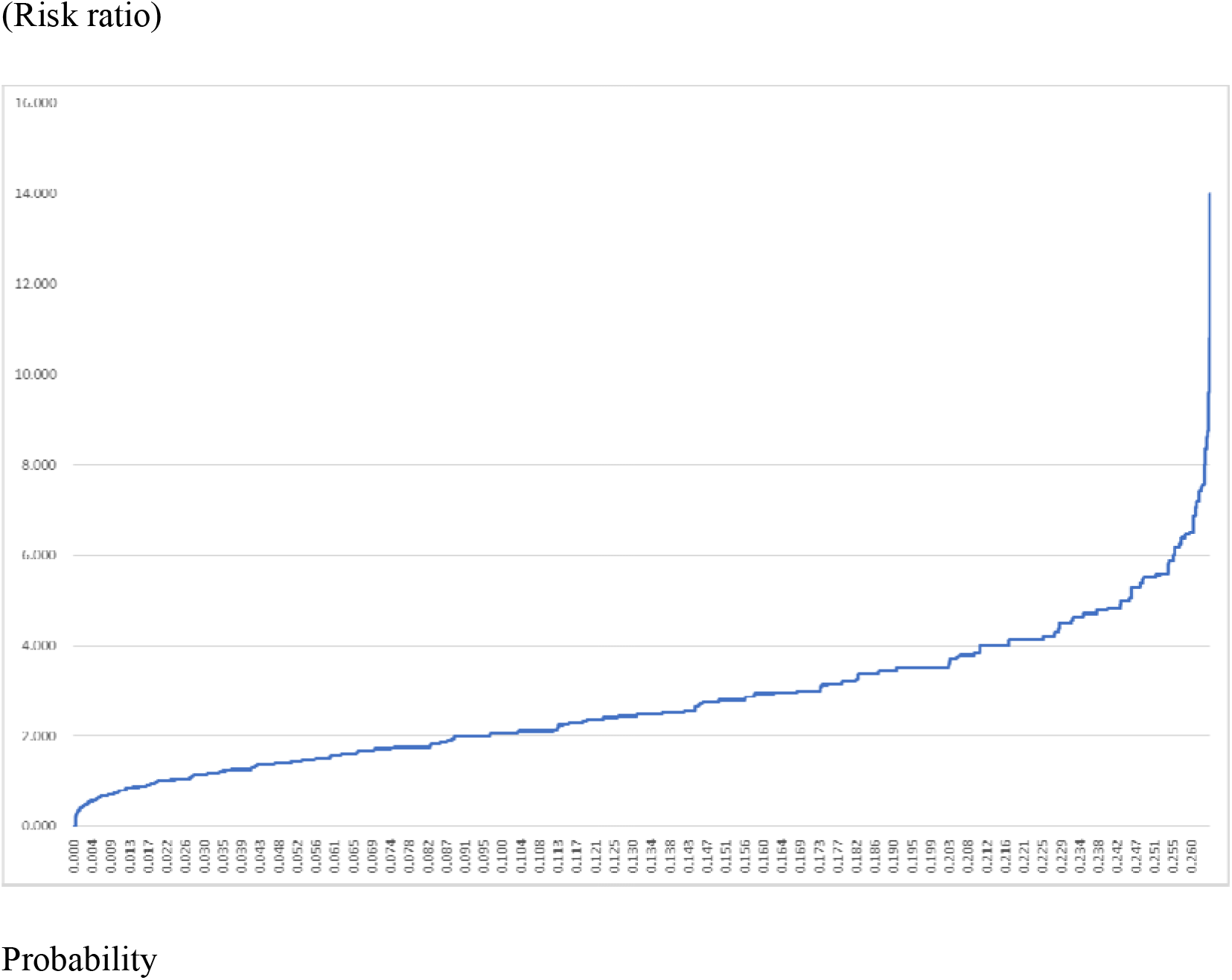
Cumulative distribution of the bootstrapped risk ratio. Note: The risk ratio is defined as the incidence in the drinking group over the incidence in the non-drinking group. Both groups had members who engaged in group dining with more than two participants and more than twice in the two weeks. If the bootstrapped incidence in the non-drinking group was zero, then the risk ratio was infinity. This figure excludes these cases.

## Discussion

Although we concluded that the null hypothesis cannot be rejected, if adopting 5% as the significance level, as done for many epidemiological studies, then we can reject the null hypothesis because the risk ratio is larger than that at 2.5%. In other words, the conclusion from this study might be affected strongly by the adopted significance level. At least one can say that high risks posed by drinking might not be supportable by strong evidence. The level of significance used to draw conclusions is determined entirely by researchers’ admissible level of error. Therefore, results must be evaluated more carefully.

The present study has some limitations. First, age, gender, and other factors that might affect behavior cannot be controlled because we have no information other than that published and used for analyses, although an earlier study [2] included control of them. Drinking behaviors might be biased according to age and gender. These tendencies might be correlated to COVID-19 infection incidence. If so, drinking itself might not present a risk of infection.

Secondly, the original data used for an earlier study [2] might include some information to test the risk of group dining with more than two participants once in the prior two weeks compared with no group dining. Alternatively, it might test the risk of group dining with more than two participants once in the last two weeks compared with group dining with one participant once in the last two weeks. These analyses might contribute to evaluation of the risks posed by the frequency and number of participants aside from those posed by drinking. We have no original data. Therefore, we cannot conduct such a study.

Thirdly, variant strain N501Y was prevalent during the study period, especially during the late half of the period. Its prevalence might affect the results of increasing risks of non-drinking.

## Conclusion

Results of this study demonstrated that no clear evidence exists to elucidate risks of drinking. However, further analyses including data collection are expected to be necessary. Policies necessitating restriction of freedoms such as restrictions against going out or drinking bans must be based on strong evidence to reduce infection effectively. Additionally, risks posed by the frequency and number of participants during group dining and drinking alcohol should be evaluated.

The present study is based on the authors’ opinions: it does not reflect any stance or policy of their professionally affiliated bodies.

## Data Availability

Japan Ministry of Health, Labour and Welfare. Documentation in the 48th Advisory Board Meeting.(in Japanese)

https://www.mhlw.go.jp/stf/seisakunitsuite/bunya/0000121431_00256.html

## Acknowledgments

We acknowledge the great efforts of all staff at public health centers, medical institutions, and other facilities who are fighting the spread and destruction associated with COVID-19.

## Ethical considerations

All information used for this study was published data on the web[2]. There is therefore no ethical issue related to this study.

## Competing Interest

No author has any conflict of interest, financial or otherwise, to declare in relation to this study.

## Notes

### Competing Interest Statement

The authors have declared no competing interest.

### Funding Statement

The authors received no specific funding for this work.

### Author Declarations

All information used for this study has been published. There is therefore no ethical issue related to this study.

## Reference

1. Post RAJ, Regis M, Zhan Z, van den Heuvel ER. How did governmental interventions affect the spread of COVID-19 in European countries ? BMC Public Health 2021 ;21:411.

2. Japan Ministry of Health, Labour and Welfare. Documentation in the 48th Advisory Board Meeting. https://www.mhlw.go.jp/stf/seisakunitsuite/bunya/0000121431_00256.html (in Japanese) [accessed on July 13, 2021].

3. Davison AC, Hinkley DV. Bootstrap Methods and Their Application, Cambridge University Press: Cambridge, 1997.

4. Mooney, .Z and Duval RD, Bootstrapping: A Nnparametric Approach to Statistical Inference. Newbury Park, CA: Sage Publication, 1993.

5. Efron B, Tibshirani RJ. An Introduction to the Bootstrap, Chapman & Hall: New York, 1993.

